# Identifying the barriers and facilitators to culturally responsive HIV and PrEP screening for racial, ethnic, sexual, and gender minoritized patients: A scoping review protocol

**DOI:** 10.1101/2023.01.18.23284716

**Authors:** Julia Xavier, Patrick G. Corr, Maranda C. Ward, Nikhil Kalita, Paige McDonald

## Abstract

**Introduction:** While mainstream messaging about human immunodeficiency virus (HIV) disparities continues to highlight individual risk-taking behavior among historically marginalized groups, including racial, ethnic, sexual, and gender minoritized patients, the effect of structural factors and social determinants of health (SDOH) on morbidity and mortality remain underestimated. Systemic barriers, including a failure of adequate and acceptable screening, play a significant role in the disparate rates of disease. Primary care practitioner (PCP) competency in culturally responsive screening practices is key to reducing the impact of structural factors on HIV rates and outcomes. To address this issue, a scoping review will be performed to inform the development of a training series and social marketing campaign to improve the competency of PCPs in this area.

**Objectives:** This scoping review aims to analyze what recent literature identify as facilitators and barriers of culturally responsive HIV and pre-exposure prophylaxis (PrEP) screening practices for historically marginalized populations, specifically racial, ethnic, sexual, and gender minoritized groups. A secondary aim is to identify themes and gaps in the literature to help guide future opportunities for research.

**Methods:** This scoping review will be performed following the framework set forth by Arksey and O’Malley and the Preferred Reporting Items for Systematic Reviews and Meta-Analyses extension for scoping reviews (PRISMA-ScR). Relevant studies between the years 2019-2022 will be identified using a rigorous search strategy across four databases: MEDLINE (via PubMed), Scopus, Cochrane (CENTRAL; via Wiley), and CINAHL (via EBSCO), using Boolean and Medical Subject Headings (MeSH) search terms. Studies will be uploaded to the data extraction tool Covidence to remove duplicates and perform a title/abstract screening, followed by a full-text screening and data extraction.

**Results:** Data will be extracted and analyzed for themes related to culturally responsive HIV and PrEP screening practices in clinical encounters with the identified target populations. Results will be reported according to PRISMA-ScR guidelines.

**Discussion:** To our knowledge, this is the first study to use scoping methods to investigate barriers and facilitators to culturally responsive HIV and PrEP screening practices for racial, ethnic, sexual, and gender minoritized populations. The limitations of this study include the analysis restrictions of a scoping review and the timeframe of this review. We anticipate that this study’s findings will interest PCPs, public health professionals, community activists, patient populations, and researchers interested in culturally responsive care. The results of this scoping review will inform a practitioner-level intervention that will support culturally sensitive quality improvement of HIV-related prevention and care for patients from minoritized groups. Additionally, the themes and gaps found during analysis will guide future avenues of research related to this topic.

## Introduction

Central to the effective care of historically excluded communities is an appreciation for cultural responsiveness, sometimes called cultural competence, among primary care practitioners (PCPs). In the context of primary care, culturally responsive communication refers to a PCP’s ability to engage with patients “based on views of culturally diverse patients rather than the views of health care professionals.”^1^ Cultural responsiveness centers unique patient experiences and understandings of health and illness, recognizes the individual biases that clinicians may hold, and seeks to work productively with patients who are not typically represented or valued in the Western understandings of care. At an organizational level, cultural responsiveness includes valuing diversity within the community; institutionalizing cultural awareness; and adapting to best serve the community by creating policies, systems, administrations, and protocols that allow for effective cross-cultural interactions.^2^ This type of approach allows healthcare practitioners to work consciously and effectively toward cultivating health equity for historically marginalized groups.

Human immunodeficiency virus (HIV) and coronavirus disease 2019 (COVID-19), for instance, are two preventable, communicable illnesses with significant burdens of disease that are highly stigmatized and disproportionately affect racial, ethnic, sexual and gender minoritized groups, hereafter referred to as “minoritized groups”. Structural factors including systemic oppression and social determinants of health (SDOH) play a significant role in the differential health outcomes of these diseases but may receive less attention than a patient’s personal factors and choices. In the landscape of these existing disparities, patients from minoritized groups are disproportionately harmed by the underutilization of culturally responsive screening practices in the primary care setting. Further, while shifts in research and funding priorities allowed for swift and large-scale responses to COVID-19, the recent pandemic has drawn much needed resources and attention away from HIV prevention and care, exacerbating the disparities of this illness and creating missed opportunities to address both diseases.^3^

To address these issues, the study team is undertaking a project to train and build the capacity of PCPs to routinize culturally responsive and nonjudgmental communication about screening and testing for minoritized groups regarding HIV and COVID-19. By providing regular, culturally responsive screening and education related to COVID-19 and HIV, PCPs can begin to bridge the divide in health disparities impacting minoritized communities. This project is multifaceted and includes data collection through scoping literature reviews and qualitative interviews that concern the experiences and beliefs of minoritized groups regarding HIV and COVID-19. These data will be used to inform and validate: a) a training series for PCPs about HIV testing, pre-exposure prophylaxis (PrEP) screening, and COVID-19 vaccine screening; b) a social marketing campaign for PCPs and by PCPs to encourage routinizing culturally responsive conversations about testing, screening, and prevention; and c) a culminating white paper on policy recommendations to improve HIV screening guidelines and to better implement existing PrEP and COVID-19 guidelines. This article discusses the protocol that will be used to perform a scoping review focused on HIV and PrEP screening, specifically.

### Background

Despite progress over the last forty years in reducing HIV transmission, morbidity, and mortality, nearly 1.2 million people are still living with HIV in the United States according to most recent data.^4^ Among those living with and becoming newly infected with HIV, minoritized groups deal with a greater burden of disease. For example, male-to-male sexual encounters made up more than two-thirds of HIV diagnoses in 2020,^4^ and a 2021 CDC special report found that 4 in 10 transgender women in major cities are living with HIV.^5^ Similarly, although Black Americans made up only 13.6% of the U.S. population in 2020, they represented more than 40% of HIV diagnoses that year.^4,6^ The same year, Hispanic or Latinx individuals made up 27% of HIV diagnoses, despite making up less than 19% of the U.S. population.^4,6^ Despite mainstream messaging that still claims these populations are more often engaged in high-risk behaviors, research data support that structural and systemic issues like socioeconomic status, housing, the physical environment, and unequal access to preventative care have contributed to these stark health disparities more significantly than individual risk behaviors.^3^

Among those living with HIV, more than 1 in 10 are not aware of their HIV status because they have not been tested.^7^ Without an understanding of their HIV status and risk, these patients may be unable to start pre- or post-exposure prophylaxis (PrEP or PEP) or the appropriate therapies to suppress their viral load. Following the current trajectory, without further intervention, close to 400,000 more people will be diagnosed with HIV by 2030.^8^ For these reasons, HIV prevention and diagnosis are central to the U.S. Department of Health and Human Services (HHS) *Ending the HIV Epidemic in the U*.*S*. plan for the next decade. The HHS plan aims to reduce new HIV infections by over 90% by 2030.^8^ PrEP coverage, a key indicator of *Ending the HIV Epidemic in the U*.*S*., shows a distinct failing. Current US Public Health Service PrEP clinical practice guidelines recommend that all sexually active patients receive information regarding PrEP and that those at substantial risk should be prescribed either daily oral PrEP or a recently approved intramuscular injection every two months.^9^ Despite this, in 2020, only 1 in 4 people in the U.S. and Puerto Rico who had indications for PrEP were receiving PrEP.^10^ Among these individuals, the aforementioned racial and ethnic disparities persist: while 66% of indicated White patients were covered, only 9% of indicated Black patients and only 16% of indicated Hispanic or Latinx patients were receiving PrEP.^10^

Moreover, while the Centers for Disease Control and Prevention (CDC) has recommended for more than 15 years that routine healthcare include opt-out HIV screening (defined as performing an HIV test) at least once for patients between the ages of 13 and 64, and at least annually for patients with risk factors,^11^ qualitative data from other arms of this study have already indicated that minoritized patients not only feel targeted and disrespected by HIV testing without context, but they report concerns that opt-out testing is or will be performed without their consent. These feelings exacerbate the mistrust that historically exploited groups have of healthcare practitioners and institutions. Even without this barrier, testing rates have not increased ubiquitously throughout the healthcare system. A 2020 Morbidity and Mortality Weekly Report by the CDC reported that HIV testing has been increasing in community health centers and emergency department settings, but not in physician offices.^12^

The above metrics support the idea that a chronic underutilization of HIV screening and prevention practices, especially in the primary care setting, misses a crucial health promotion opportunity, and disproportionately harms members of historically marginalized groups due to the systemic barriers that exist within healthcare and societal institutions. It is the responsibility of PCPs to understand these barriers and to identify the impacts of structural racism on the poor outcomes and delayed identification of HIV in minoritized groups so they may prioritize culturally responsive communication with these patient populations. The definition of HIV screening must expand to include these conversations about HIV and PrEP to conscientiously serve historically marginalized patients. Moving forward, we use the term “screening” to indicate screening conversations and communication practices among PCPs that facilitate informed HIV testing and connection to PrEP.

To begin meeting these aims and the study goals outlined above, we will conduct a scoping review to assess existing literature for insights about this topic. To our knowledge, there has not yet been a scoping review regarding this subject and the target patient populations. A preliminary search of databases identified no ongoing or existing systematic/scoping reviews on this topic. Therefore, the scoping review will be conducted with two specific purposes. Firstly, to inform our understanding of the factors which influence culturally responsive HIV and PrEP screening conversations for historically marginalized populations. The results of this review will inform the design of a training series and social marketing campaign targeted at PCPs. Secondly, the review will identify themes and gaps in the current literature to guide future research opportunities in this field.

### Timeline

This scoping review was designed in October 2022, reviewed with members of the research team, and shared with university reference librarians for initial feedback. Data extraction and analysis will begin in late January 2023 and continue through March 2023. We anticipate submitting the results of this scoping review for publications in March 2023.

### Methods

Scoping reviews differ from systematic reviews in that they focus on reviewing the breadth, rather than depth of current literature regarding a particular topic. For this reason, they may be completed much more rapidly than systematic reviews, which take time to assess the quality of evidence among studies. A scoping review was chosen to quickly ascertain the nature of the knowledge base surrounding factors that influence culturally responsive HIV and PrEP screening for patients identifying as members of minoritized groups.

The scoping review will be performed according to the framework first outlined by Arksey and O’Malley.^13^ The five steps of this process include: 1) identifying a research question, 2) identifying relevant studies, 3) selecting studies, 4) charting the data, and 5) collating, summarizing, and reporting the results. This will also be guided by the specific steps of the PRISMA extension for scoping reviews (PRISMA-ScR),^14^ several of which will be described in relevant sections below.

### Step 1: Identifying a research question

The primary research question (PRISMA-ScR Item 4: Objectives) in this review will be, “*what factors influence culturally responsive HIV and PrEP screening for historically marginalized populations?”* to explore the ways we might disrupt structures of power working against patients from minoritized groups. A secondary question will be, “*what themes and gaps exist in the literature regarding culturally responsive HIV and PrEP screening for historically marginalized populations?”* to identify areas of interest for future research.

### Step 2: Identifying relevant studies

The scoping review will be conducted across four databases (PRISMA-ScR Item 7: Information Sources): MEDLINE (PubMed), Scopus, Cochrane (CENTRAL; Wiley), and CINAHL (EBSCO). The search strategy includes the use of Medical Subject Headings (MeSH) terms and Boolean terms to clarify the parameters of the review. After conducting the searches on each database, the results will be imported into Covidence to remove duplicates and perform primary screening of titles and abstracts for relevance. The full search strategy (PRISMA-ScR Item 8: Search) is presented in Table 1.

**Table 1:**
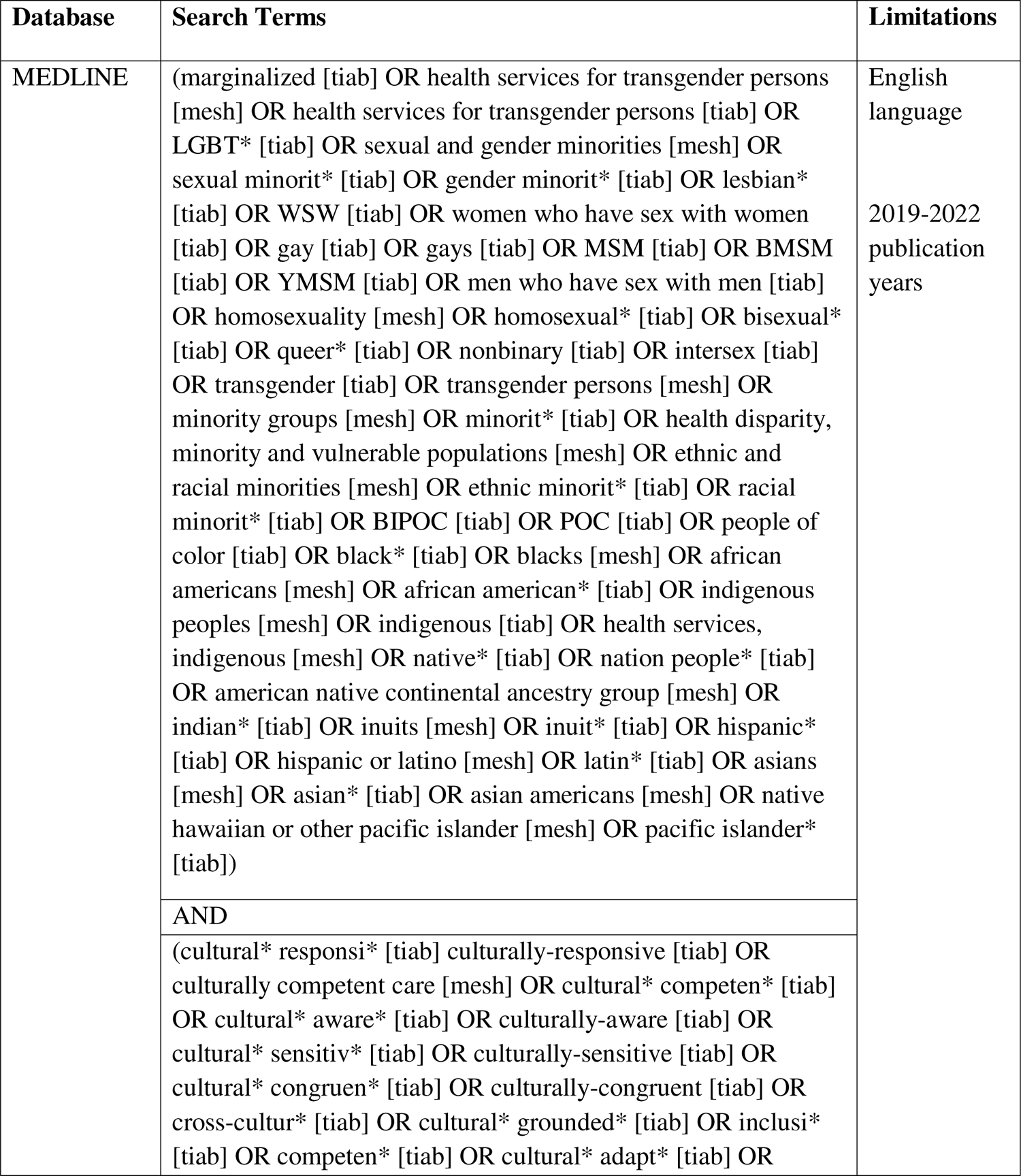
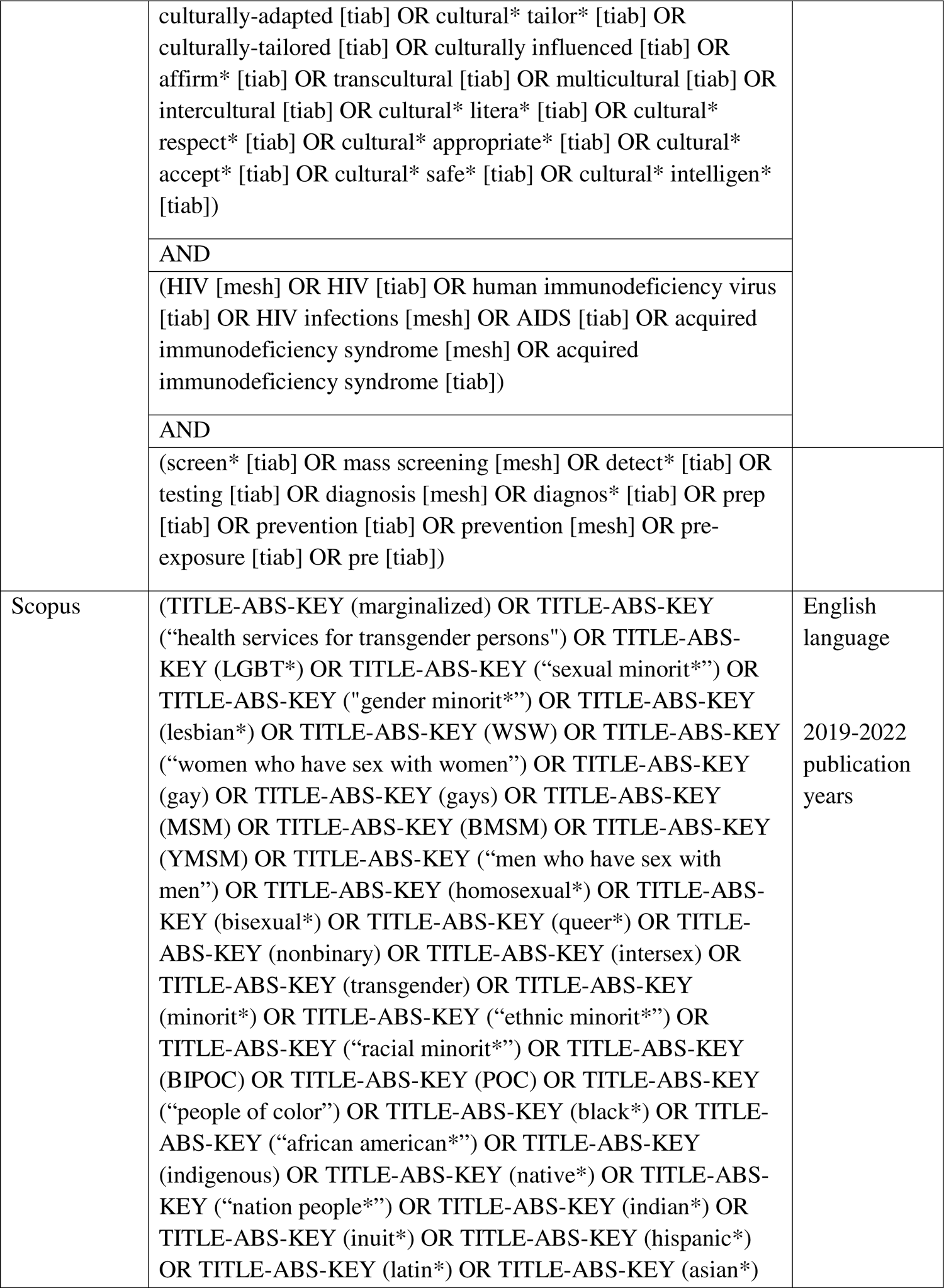
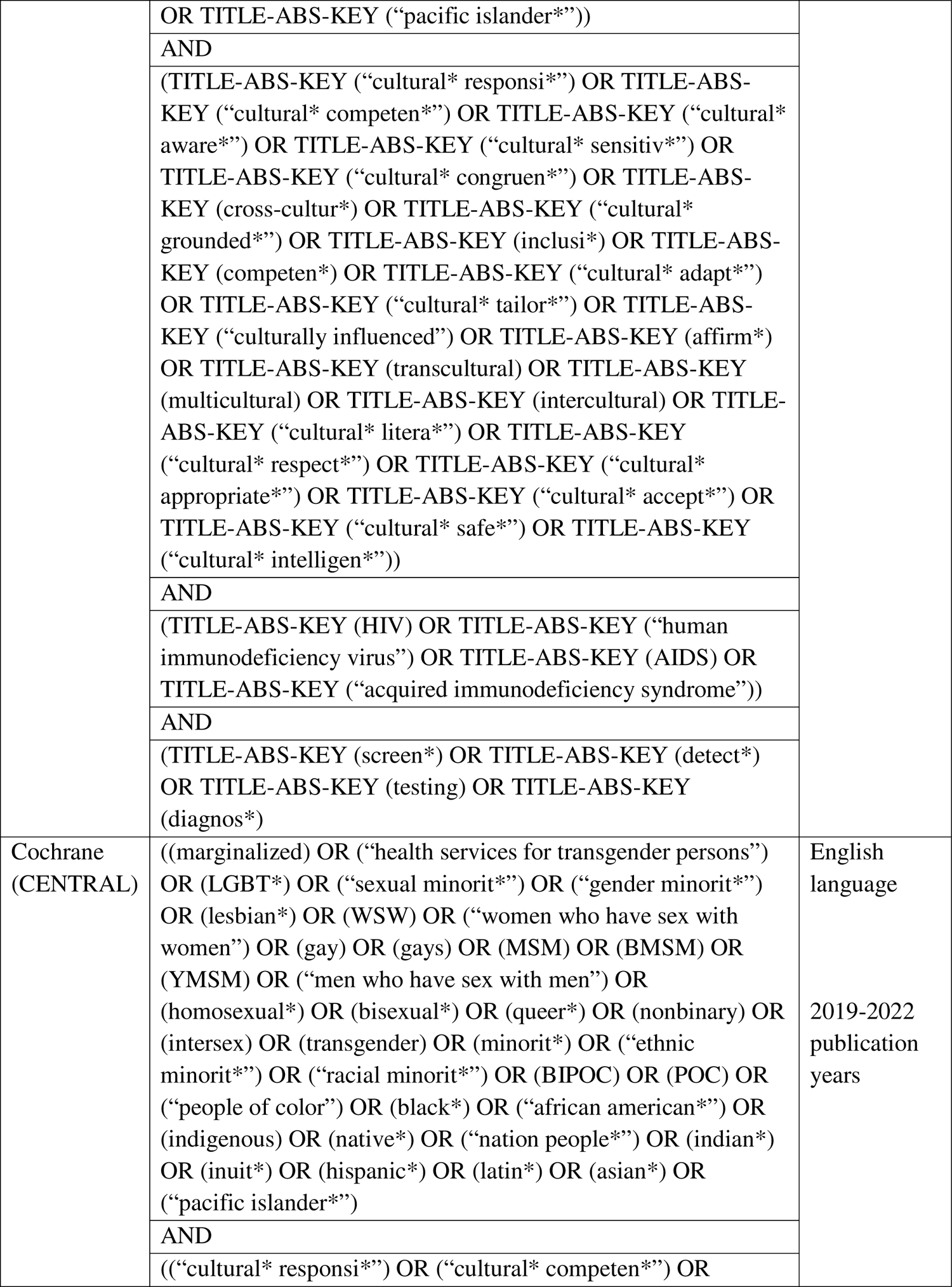
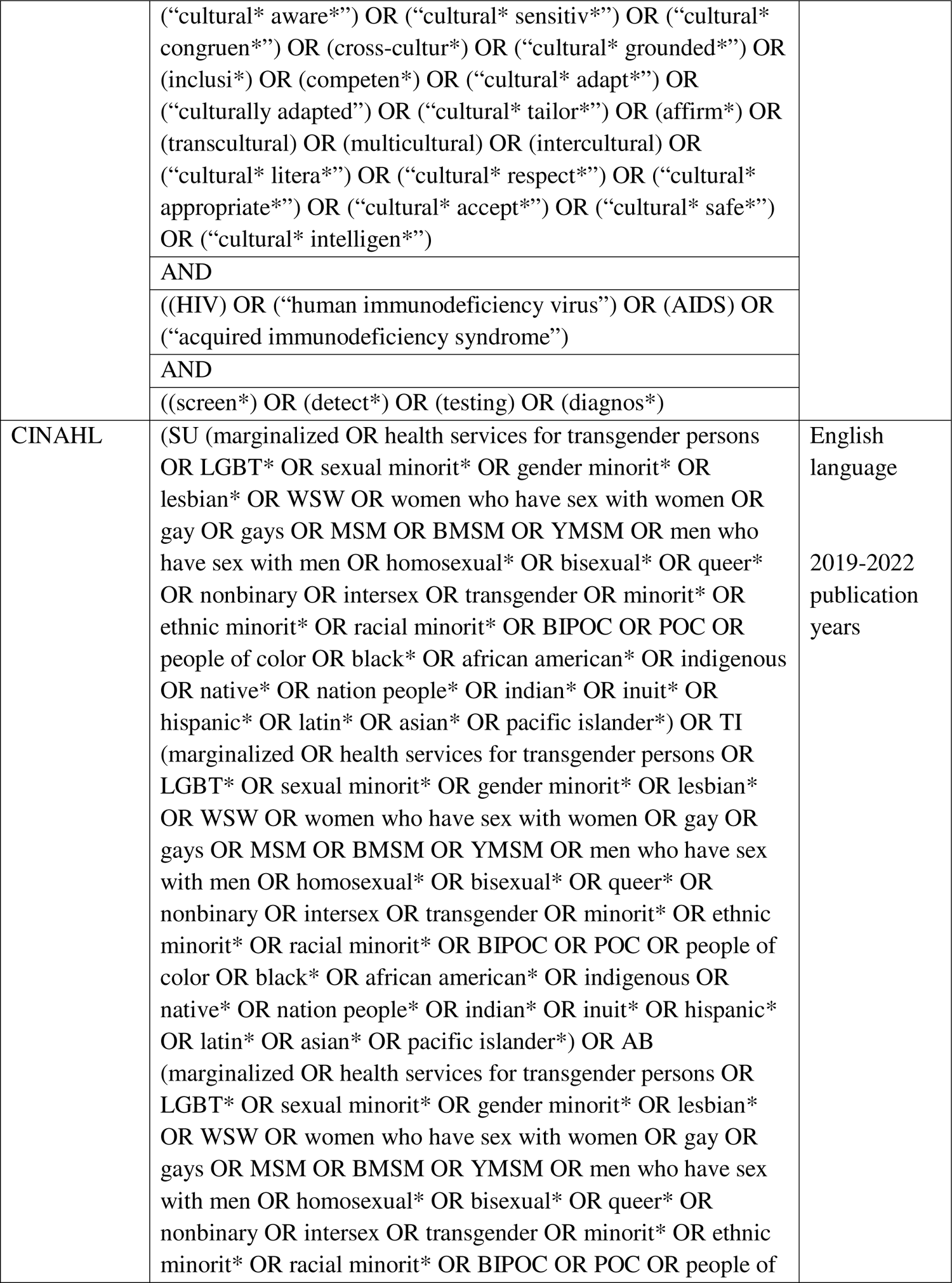
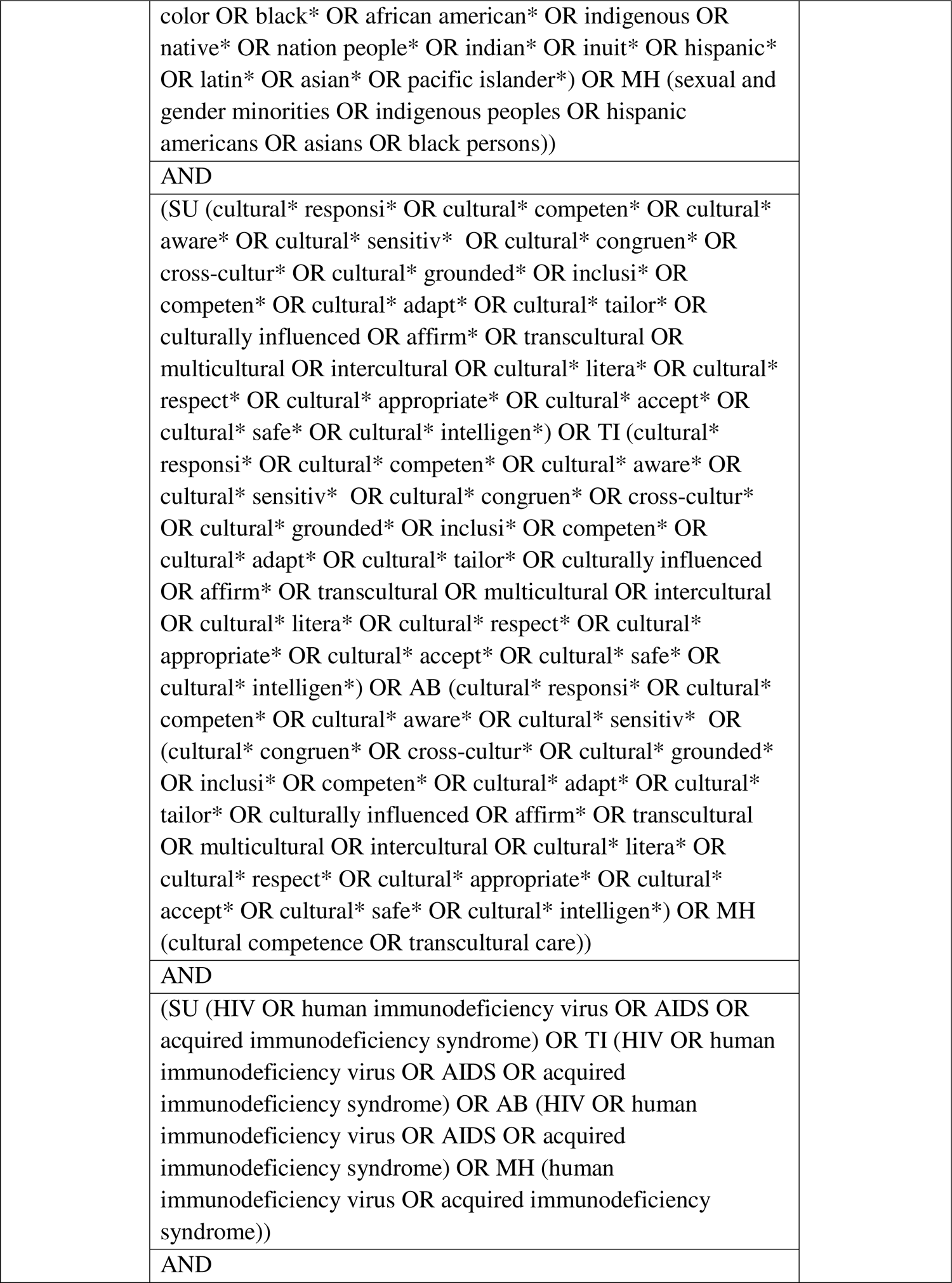
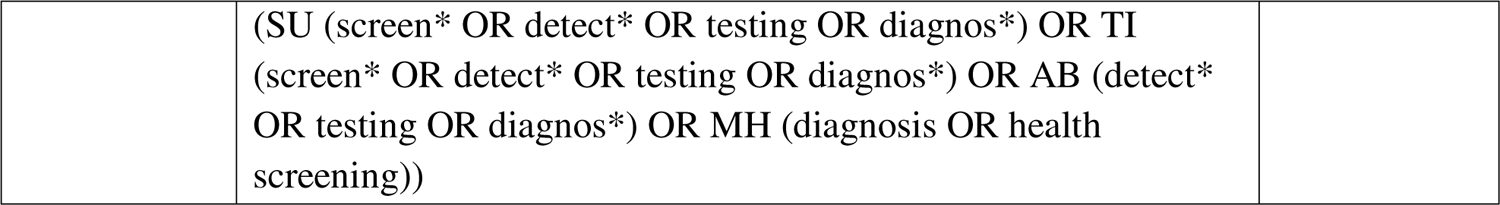
Search Terms and Limitations

### Step 3: Selecting Studies

#### Inclusion Criteria

The search will be limited (PRISMA-ScR Item 6: Eligibility Criteria) to studies that concern screening services, specifically regarding HIV and/or PrEP. In addition to the search strategy delineated above (**Table 1**), studies will be included in the review if they meet the following inclusion criteria:

⍰ Published, peer-reviewed research articles
⍰ Studies published in English
⍰ Studies that consider data collected in the U.S./in the context of U.S. health systems

#### Exclusion Criteria

Studies that meet the following exclusion criteria will be excluded from the review:

⍰ Studies where the team cannot obtain full-text articles
⍰ Book chapters and study protocols
⍰ Studies published in a language other than English
⍰ Studies published prior to 2019
⍰ Data collected from any country other than the U.S., except for systematic reviews
⍰ Studies that do not address one or more of our historically marginalized populations of interest or studies that do not discuss people of unknown or negative HIV status

Results will be limited to published, peer-reviewed studies to create a standard for included articles, as they will not be assessed for quality of evidence. Results must have been published in English, as the team does not have the resources to translate foreign language studies. Further, since our training intervention seeks to improve outcomes among patients in the U.S. by understanding barriers and facilitators among U.S.-based PCPs, results will be limited to studies that were conducted in the U.S. or that concern U.S. health systems. Study protocols will be excluded because they are not predicted to be of use, and book chapters will be excluded to avoid the use of non-peer-reviewed materials. Since the intervention ultimately informed by this scoping review is two-pronged and targets both HIV and COVID-19 screening, included studies will be limited to those published between the years 2019-2022 (mirroring the timeline of the COVID-19 pandemic), rather than including HIV-related research from the past four decades. Finally, since this review will inform interventions regarding HIV and/or PrEP screening for patients from racial, ethnic, sexual, and gender minority backgrounds, studies must focus on at least one of these groups, and must include. Articles will necessarily be limited to those that include individuals of negative or unknown HIV status, because these are the patients who are eligible to be screened for HIV and PrEP.

Results will be uploaded to Covidence, a screening and data extraction tool, to eliminate duplicate studies, followed by two stages of screening: a title/abstract screening, followed by a full-text screening (PRISMA-ScR Item 9: Selection of Sources of Evidence). The primary reviewers (JX, NK) will screen all the results against the inclusion and exclusion criteria and must agree to include or exclude a study to make a decision. Senior reviewers (PM and PC) will resolve disagreements. After completing the title/abstract screening, reviewers (JX, NK, SP, DB) will perform a full-text screening on the remaining studies and further eliminate irrelevant studies, and PM and PC will provide guidance and reconciliation as required. To assess interrater reliability, all reviewers will informally screen the same several articles at the start of both title/abstract screening and full-text screening phases and come together to discuss the criteria/process used to include or exclude studies.

### Step 4: Charting the data

After studies have undergone a full-text screening, data will be extracted and charted by nine reviewers (JX, NK, SP, DB, SS, PC, AK, MW, PM) (PRISMA-ScR Item 10: Data Charting Process). The data items for extraction (PRISMA-ScR Item 11: Data Items) were developed through an iterative process that consulted team members and created a working definition of each item to streamline the extraction process. Checkboxes were chosen over text fields for several items to minimize variability among different reviewer inputs. The data that will be charted includes:

⍰ Reviewer
⍰ Covidence ID
⍰ Author(s)
⍰ Publication title
⍰ Publication year
⍰ Study location
⍰ Study design
⍰ Intervention type
⍰ Timeframe of study
⍰ Study aims
⍰ Study population
⍰ Methodology overview
⍰ Measures
⍰ Results
⍰ Level of communication addressed
⍰ How does it address Black Indigenous, and People of Color (BIPOC) groups?
⍰ How does it address lesbian, gay, bisexual, transgender, queer/questioning, intersex, and asexual (LGBTQIA+) groups?
⍰ What aspects of cultural responsiveness are addressed?
⍰ Facilitators to culturally responsive communication identified
⍰ Barriers to culturally responsive communication identified
⍰ Implications
⍰ What is left unanswered?
⍰ Limitations or biases

## Results

### Step 5: Collating, summarizing, and reporting the results

After extraction, reviewers will quantify mentions of specific barriers and facilitators to identify their prevalence, then pull quotations from each included study to describe each identified influence. The implications extracted from each study will be analyzed for themes related to culturally responsive HIV and PrEP screening practices for historically marginalized populations. The data will be reviewed to identify how each study contributes to our understanding of the primary research question. Additionally, the review will identify any gaps in existing literature and opportunities for future research in this field. This analysis will not include a systematic evaluation of study quality or risk of bias, because this scoping review aims to focus on the scope and breadth, rather than the quality of available literature. After analysis, results will be synthesized and reported according to PRISMA-ScR guidelines (Item 13: Synthesis of Results; Items 15-19), and the process used to select studies will be detailed in a PRISMA flowchart (PRISMA-ScR Item 14: Selection of Sources of Evidence).

## Discussion

Culturally responsive HIV screening conversations between PCPs and minoritized groups are key to bridging the stark existing disparities in HIV morbidity and mortality in the U.S. To our knowledge, this is the first study to use scoping methods to investigate the barriers and facilitators to culturally responsive HIV and PrEP screening for minoritized populations. After mapping the landscape, gaps, and themes of current research, we plan to disseminate our findings in publication and through the development of relevant and evidence-based training programs to help PCPs routinize culturally responsive HIV and PrEP screening for minoritized groups. Additionally, we anticipate that this study will yield implications important to the work of activists, researchers, patients, and public health professionals in this field. The limitations of this study include that as a scoping review, our review does not assess the depth, quality, or risk of bias of the studies the way a systematic review would. Additionally, by limiting the timeline of this review to the years 2019-2022, we may miss valuable studies that were performed before this timeframe.

## Data Availability

No datasets were generated or analysed during the current study. All relevant data from this study will be made available upon study completion.

## Institutional Review Board Statement

This project did not utilize human subjects, was exempted from needing IRB approval, and therefore did not involve a process of informed consent. This manuscript exclusively provides an overview of a scoping review protocol. The data from this scoping review will be used to inform and strengthen an educational intervention that will be IRB-approved and supported by grant funding.

## Acknowledgments

The authors would like to acknowledge Thomas Harrod, Associate Director of Reference, Instruction, and Access at the George Washington University’s Himmelfarb Health Sciences Library for his generous guidance and support in developing the search strategies for this scoping review and Stacy Brody, Reference and Instruction Librarian at the George Washington University’s Himmelfarb Health Sciences Library for her guidance in creating the manuscript for this publication. The authors would like to acknowledge Saylor Pershing, Darrell Bailey, Sheel Singh, and Abigail Konopasky for their assistance in the text screening and extraction process.

